# Engaging Private Pharmacies in Referring Tuberculosis Suspects in Pakistan: Findings and Implications

**DOI:** 10.1101/2025.01.14.25320381

**Authors:** Zikria Saleem, Ayyaz Kiani

## Abstract

**Background:** World Health Organization (WHO) considers local pharmacies as an underused source that can contribute more effectively to the health care programs of the community. This study was aimed to evaluate the impact of involving private pharmacies for timely referral to facilitate TB case detection in Pakistan.

**Methods:** In this study, 398 pharmacies in five major districts of Pakistan were contacted. Patients having apparent TB symptoms or chronic cough for more than two weeks were referred for TB case detection to the nearby center of National TB Control Program (NTP). The patients were monitored and followed up to analyze the results of the diagnostic tests by involving pharmacy students.

**Results:** Out of 398 enrolled pharmacies, only 224 pharmacies (56.28%) were producing referrals. A total of 994 TB suspects were referred for tests out of which 839 (84.4%) actually appeared for the TB examination tests while 155 (15.5%) were missing. Of total 839 tested suspects, 189 (22.5%) were diagnosed with smear positive TB.

**Conclusion:** Community pharmacies under the thematic model of public private partnership can be the key players in tracing, preventing and early diagnosis of TB. This project can be extended and implemented in all provinces of Pakistan with more technical and financial inputs in the existing system in order to completely eradicate TB.

## 1. INTRODUCTION

Strengthening health care systems to make them more effective in meeting the needs of the population is currently the main target of governments in many developing countries[1,2]. World Health Organization (WHO) considers local pharmacies as an underused source that can contribute more effectively to the health care programs of the community [3,4]. They can also play a major role in increasing the diagnosis of wide spread communicable diseases like TB (TB). TB is one the of the leading death-causing diseases around the world despite the availability of cheap medicines and effective treatment[5]. Poorest persons in the low to middle income countries suffer the most from this disease[6]. However, improved results in TB treatment have been shown in the recent past due to the developments in diagnostics and medicine[7]. The treatment success rate among new TB cases are improving but there is a need of major efforts to ensure that all the TB cases are detected on time, notified and treated [8,9]. Community pharmacies have quite a distinct and unique position in the healthcare delivery system as they are the first contact for patient getting medicines in the majority of cases [10,11]. Their importance is enhanced by their large coverage and magnitude of operations, serving millions of patients every day [4]. However, an important fact to consider is that these outlets are working mainly as business entities and not as healthcare providing institutions especially in low and middle income countries [12-14].

TB is a curable and preventable bacterial infection caused by *Mycobacterium TB* that affects the lungs, bones, intestine and other organs. In 1993, the TB cases were increasingly reported worldwide, compelling the WHO to declare it as a global emergency [15]. The patient with active TB is the source of dissemination of bacteria in the air through cough, sneeze or spit and is the main cause of spread. Almost one third of population of the world has either TB infection or they are active TB patients [5]. In 2013, 9 million people were affected from TB and 1.5 million died due to this disease [16]. According to a study, 95% TB patients belongs to low to middle income countries [9]. TB is also associated with Human Immunodeficiency Virus(HIV) and it kills one fourth of the patients that are infected by HIV[17]. According to Global TB Report 2014, an estimated 3.5% and 20.5% cases of newly diagnosed and previously treated multidrug resistant TB (MDR-TB) cases have been reported respectively. About 480,000 new incident cases were reported and 210,000 people died due to MDR-TB in 2013[18]. The TB death rate is dropping as compared to previous decades but the goal of eradicating TB is still far from being achieved [19].

The Sustainable Development Goals of the United Nations focus on reducing the incidence of TB [20].The WHO recommends the Stop TB Strategy based on Directly Observed Treatment Short-Course(DOTS) to control TB [21]. The existing control strategies lack the improved methods of TB detection and early identification of multidrug-resistant TB[22]. In the list of TB high-burden countries worldwide, Pakistan stands at fifth rank [9]. Around420,000 new TB cases emerge every year and almost half of them are smear positive which makes Pakistan 4^th^ among multi-drug resistant TB (MDR-TB) countries [9,16,23,24]. Government run programs in the low to middle income countries that are modest in cost are quite successful in providing high quality health care to the public. In this context private run pharmacies can improve diagnosis and treatment. A study from Karachi, Pakistan indicated that involvement of private healthcare settings as National TB Program (NTP) reporting centres, can help in early detection of TB via timely referral of the patients [25]. Another study from Sindh, Pakistan reported that only 14% of General Practitioners (GPs) performed sputum test among patients, indicating a strong need of partnership between GPs and National TB Control Programs (NTP) to improve case detection and treatment strategies [26]. A study involving public private mix model towards eradication of TB from Karachi, Pakistan indicated that 87% of the enrolled patients completed their therapy successfully where time, cost and effort to implement the TB control via public private partnership was much higher than many other countries [27].Involving private pharmacies along with the government run programs are useful for two strategic reasons; firstly to improve the TB management practices of private pharmacies and secondly to improve the access of people to good TB care [28]. The current study aimed to provide timely referral of patients for TB case detection and to evaluate the impact of involving private pharmacies as an initiative of PPM for timely referral to facilitate TB case detection in Pakistan.

## 2. METHOD

### 2.1 Study Site

The study was conducted in five major cities of Pakistan. The cities included were Lahore, Islamabad, Peshawar, Rawalpindi and Sukhur. Lahore and Peshawar are provincial capitals while Islamabad is the federal capital of the country. After developing a sampling frame, total number of 398 pharmacies and medical stores from five cities of Pakistan were selected for the study by using non probability quota sampling technique. Pharmacies and medical stores were stratified according to the selected locations. Pharmacies and medical stores inclusion criteria included the premises of the selected catchment area having license issued by the Director General Health Services, Department of Health, Government of the Pakistan. Exclusion criteria were those pharmacies and medical stores that sold only to wholesale distributors and those located within public hospitals. Before participating in this private public mix programme, pharmacy and medical store owners or staff attended training for a period of one week, carried out by pharmacy schools of respective city. Pharmacy and medical store staff were instructed to recognize, encourage and pass on TB indicative customers presenting at the medical store and pharmacy. Medical store and pharmacy owners and personnel who completed the training were the nominated TB referral service provider. Medical stores and pharmacies were given no imbursement for rendering TB referral services.

### 2.2 Study population

Study population comprised of TB suspects that were identified and sent to the TB DOTS centers for tests. Walk in customers of all age groups with any three of the following symptoms related to TB-cough for more than 2 weeks, loss of appetite, weight reduction, blood in sputum, fatigue, and fever at night-were included in the study. Customers with previous history of TB were also re-evaluated through this study. Customers who were in direct contact of a confirmed TB patient were also included.

### 2.3 Ethical Approval

A Memorandum of understanding (MOU) was signed with the universities and pharmacies which had relevant information regarding the project. The information included objectives of the study, its beneficial health outcomes such as timely referral leading to reduced disease burden and how the project will be conducted. Informed consents were obtained from all the participants of the study (who were randomly enrolled from the pharmacies). The study was regularly monitored and assessed throughout the program by project supervisors. The ethics approval was taken from the Ethics Committee on Human Research of University College of Pharmacy, University of Punjab with reference number HEC/PUCP/1953.

### 2.4 Tools of measure and data collection

Referral slip booklets were distributed among all the pharmacies that agreed to work under the Public-Private mix (PPM) TB Referral program by signing Memorandum of Understanding (MOU). The referral slips included all the information regarding the patient and the center which is recommended for the tests (Figure 1).Fifteen to twenty Pharm. D students of final professional year were trained for the program in each city. Students were divided into teams and pharmacies were allocated to them in different sites and locations of the cities. The task of the students was to collect referral slips of the patients from the pharmacies and to get follow up of the suspected TB patients. The pharmacists, pharmacy technicians, salespersons and owners of the pharmacies and medical stores were also trained and demonstrations and mock exercises about the referral processes of TB were done. All the referred patients were given a referral slip for a nearby TB diagnostic center or hospital. The patients who had any of the symptoms mentioned on referral slip (Figure 1) were referred for screening. The diagnosis was based on Referring symptomatic customers to their neighboring or most well-situated health center aims to curtail transport expenses for customers and assist their well-timed appearance at the health center for TB diagnosis. Smear-positive patients reflected the presence of TB. Additionally, X-ray analysis was performed by the clinical staff including the physicians and laboratory staff to confirm the diagnosis. The referred suspects were tested for TB free of cost and all the suspects which were diagnosed with TB were given free treatment by the National TB Control Program (NTP) upon showing the referral slip.

**Figure 1:**
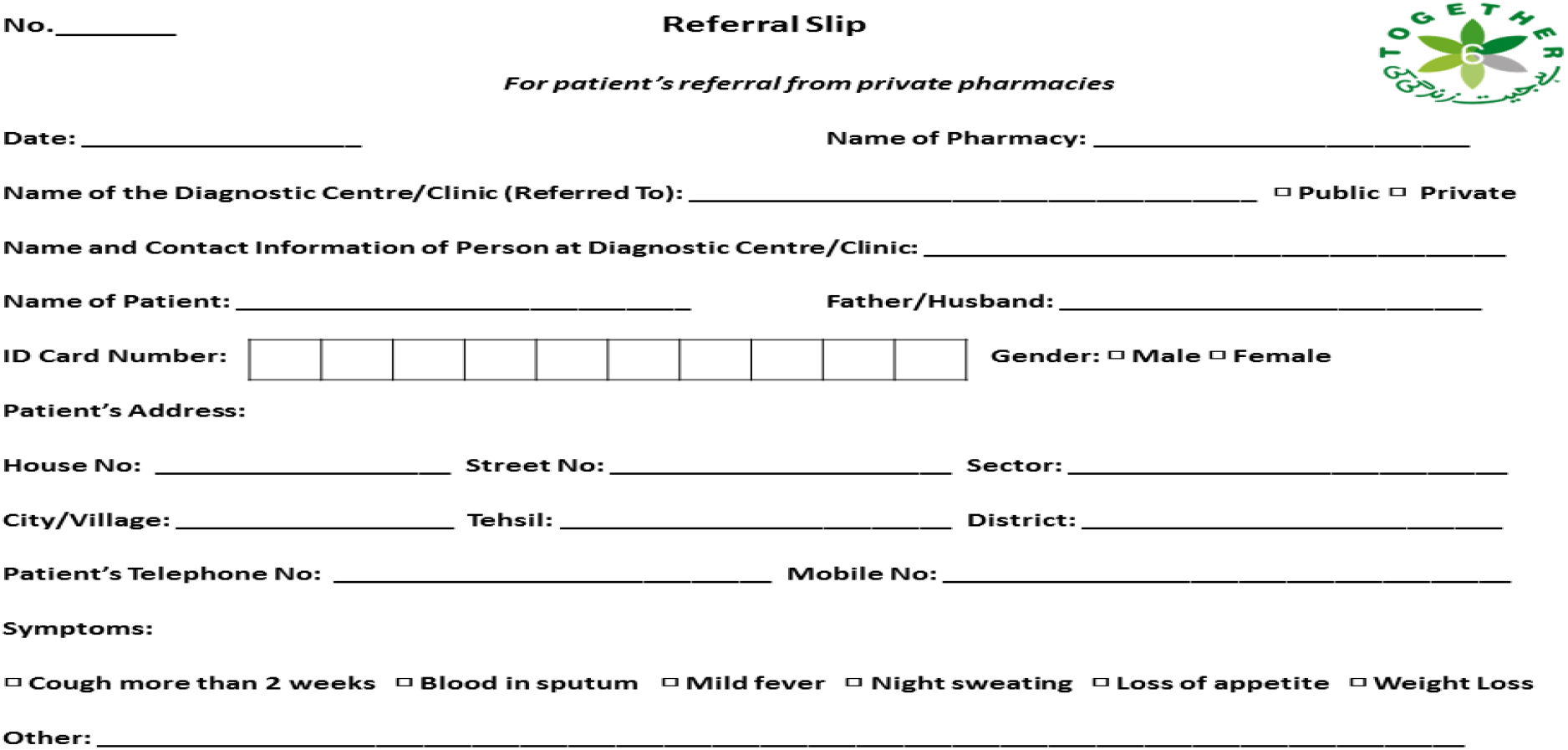
Structure of Referral slip used for Screening & Follow up

### 2.5 Monitoring and Evaluation System design, validation and implementaton

The Study model was based on public private partnership under National TB Control Program, therefore, the number of referrals and visits to the pharmacies were recorded through online tracking using Google Sheets. Tracking sheets were updated on daily, weekly, fortnightly and monthly basis, according to the status of referred patients by the respective pharmacies. The whole study was carried out for a period of 8 months, starting from December 2014 to July 2015.

### 2.6 Data analysis

The data was analyzed using statistical program for social sciences (SPSS).Descriptive statistics were applied where simple frequencies were used to present the referral, testing and detection from different districts of Pakistan.

## 3. RESULTS

Based on the referral slips and monitoring and evaluation system designed, a total of 994 patients were referred from 398 pharmacies in 7 months (Table 1& Figure 2). These patients were referred for screening to the hospitals and clinics affiliated with National TB control program (NTP). The status of 155 (15.5%) patients was missing due to unavailability of contact numbers; therefore the follow up for those patients was not possible. A total of 839 patients (84.4% of the total referrals) went for the screening test after being referred to the TB diagnostic centers. The result of the tests of 350 (41.7 % of the tested patients) out of those 839 patients was found to be negative. There was no response from 300 patients due to unknown reasons/circumstances. A total of 189 patients were smear-positive TB. Most of smear positive cases were identified from Rawalpindi (46%) where on an average 7.4 positive cases were identified by 10 pharmacies. The number of referrals and subsequently smear positive TB cases gradually increased over the period of time as the pharmacists got familiar with the program and the process. Out of 398 enrolled pharmacies, only 224 pharmacies (56.28%) were producing referrals. Among all districts, only 44% pharmacies of Lahore city were referring the patients to TB Centers. On an average; nearly 5 (3.2-7.4) new TB cases were identified as per 10 enrolled pharmacies.

**Table 1:**
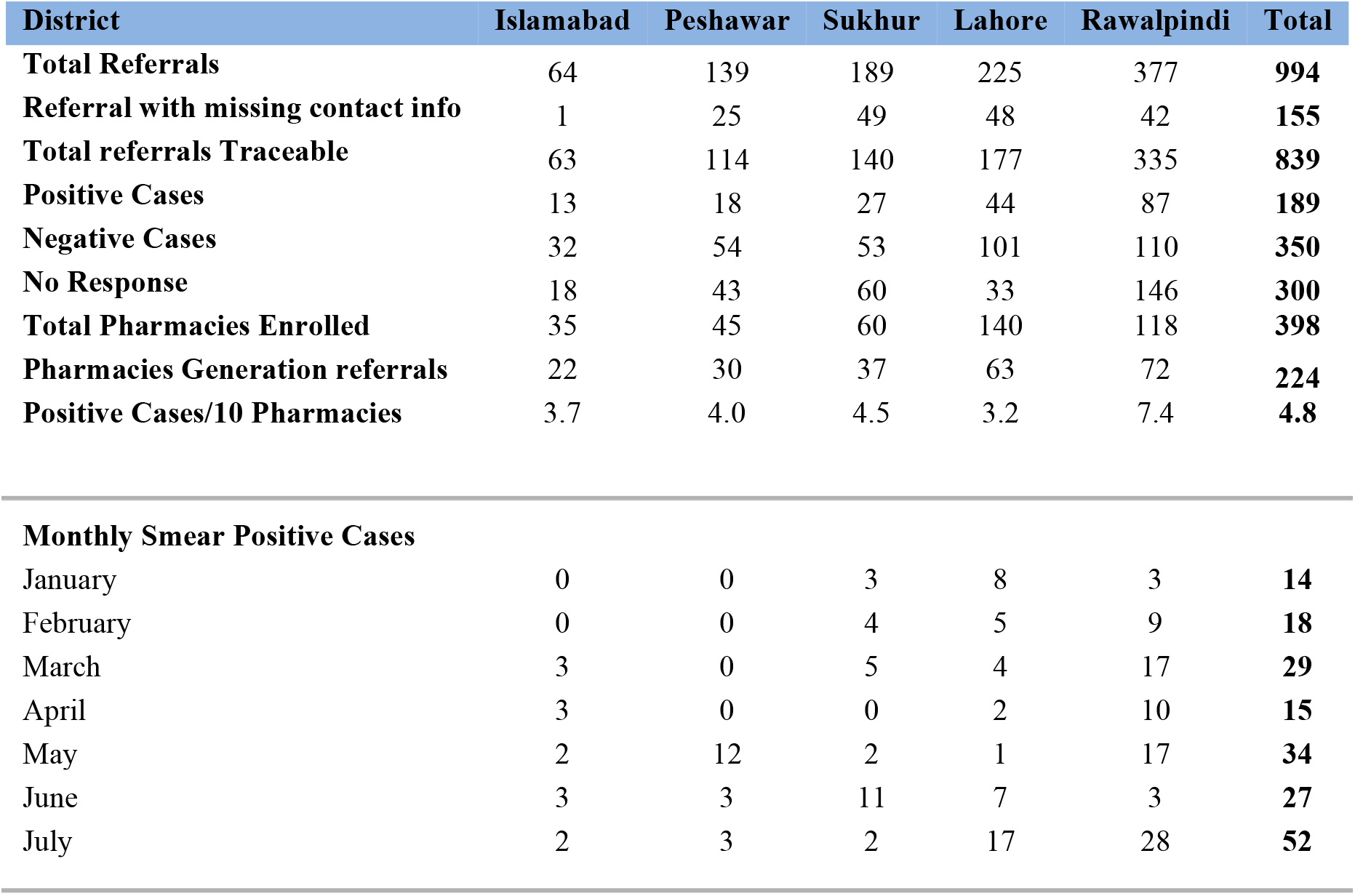
Referring, testing and detecting rates of sputum smear-positive cases of referred clients from different districts in Pakistan.

| District | Islamabad | Peshawar | Sukkur | Lahore | Rawalpindi | Total |
| --- | --- | --- | --- | --- | --- | --- |
| **Total Referrals** | 64 | 139 | 189 | 225 | 377 | **994** |
| **Referral with missing contact info** | 1 | 25 | 49 | 48 | 42 | **155** |
| **Total referrals Traceable** | 63 | 114 | 140 | 177 | 335 | **839** |
| **Positive Cases** | 13 | 18 | 27 | 44 | 87 | **189** |
| **Negative Cases** | 32 | 54 | 53 | 101 | 110 | **350** |
| **No Response** | 18 | 43 | 60 | 33 | 146 | **300** |
| **Total Pharmacies Enrolled** | 35 | 45 | 60 | 140 | 118 | **398** |
| **Pharmacies Generation referrals** | 22 | 30 | 37 | 63 | 72 | **224** |
| **Positive Cases/10 Pharmacies** | 3.7 | 4.0 | 4.5 | 3.2 | 7.4 | **4.8** |
| **Monthly Smear Positive Cases** |  |  |  |  |  |  |
| January | 0 | 0 | 3 | 8 | 3 | **14** |
| February | 0 | 0 | 4 | 5 | 9 | **18** |
| March | 3 | 0 | 5 | 4 | 17 | **29** |
| April | 3 | 0 | 0 | 2 | 10 | **15** |
| May | 2 | 12 | 2 | 1 | 17 | **34** |
| June | 3 | 3 | 11 | 7 | 3 | **27** |
| July | 2 | 3 | 2 | 17 | 28 | **52** |

**Figure 2:**
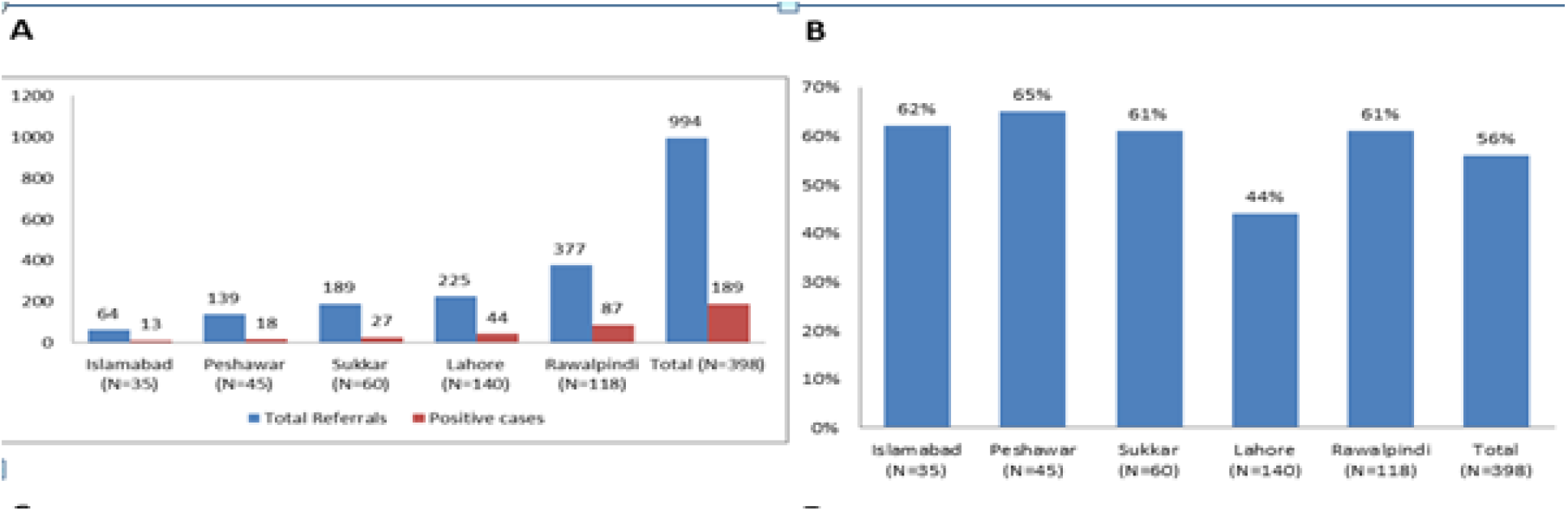
Referring, testing and detecting rates **(A)** Comparison of total referrals vs. sputum smear-positive cases **(B)** Percentage of referrals from different districts

## 4. DISCUSSION

The results showed that the project was a success in Pakistan as compared to the other similar project conducted in Cambodia [29]. The PPM model for control of TB by the intervention of the private pharmacies to increase case detection is a new method to be used on such a scale in Pakistan, although this PPM program has been conducted in different capacities in various countries of the world as Vietnam, Bolivia, India, Nepal, Indonesia, Kenya and Egypt[30]. By involving private physicians, it has also been launched in Thatta, Sindh in 2007 which was the first of its kind in Pakistan for early case detection [31]. In our study, out of 839 tested patients, 22.7 % (n =189) patients were detected with smear positive TB and they were provided with free treatment. This was estimated to be a great success considering the current scenario of health in Pakistan [31]. Contrary to the opinion of the authors of a similar program in Bolivia, who concluded that efforts and funds needed to increase the TB case detection should be invested somewhere else instead of this sort of program, [32] we believe that this result of 189 positive TB cases detected as a result of intervention of private pharmacies and medical stores is a good achievement. A similar intervention was also conducted in Vietnam that involved monitoring of 150 pharmacies over 9 months. The study in Vietnam contributed 1% increase in total case detection of smear-positive TB in the National TB program [33]. Similar results were concluded by Public Private Mix study in India (Pune & New Delhi) and Kenya [28]. The percentage of case detection in Pakistan was almost three times more than the other countries in which such project has been conducted [30]. Many difficulties were faced in running the project but the success of 994 suspects sent to diagnostic centers for TB tests and getting 189 positive cases detected will help to develop this type of projects on a larger scale in context of the control of TB in Pakistan. The difficulties faced on behalf of the suspects included lack of time, lack of trust on these kind of programs, disbelieve among the suspects related to experimentation associated with their health and fear of being diagnosed with tuberculosis. Further, suspects prefer consulting their family physicians. Other difficulties included lack of cooperation by the pharmacists and other pharmacy staff members due to the fear that customer may be neglected and will switch to other pharmacies to purchase the medicine. The main advantage of this project was for the patients who could get tested for free and got free medications as well. They utilized the opportunity of referral slips to get themselves tested and also utilized this program to get quality treatment for the cure of TB. Finally, this project not only improved the detection of the undiagnosed TB patients in the city but also screened the problems that can be sorted out to make this project successful on a larger scale.

Engagement of the private sector pharmacies and medical stores is a recommended strategy for increasing TB case detection and managing treatment[33,34]. Involvement of all the sectors related to health is necessary to prevent, detect and treat the patients of TB. Efforts were started with the involvement of physicians but now the whole health community including the pharmacists and other organizations related to health are included in the aim to prevent, treat and diagnose the cases[35]. Pharmacies and medical stores are believed to be an obstacle in the treatment of TB globally because they often dispense palliative medication, mono-therapies, insufficient dosage and a source of self-medication that prove to be a hindrance in the treatment and also results in non-compliance of the patient to the treatment[36,37]. However, efforts to promote referrals by pharmacies and medical stores in some countries of the world have shown success in increasing the case detection of the patient[29-31]. Timely referrals have increased and it has also helped in reducing the unregulated sales of TB medicines[29]. Consequently, pharmacies are playing an important part in early case detection of TB patients in many countries of the world [34]. Public-private mix partnership is believed to be necessary for countries with high prevalence of TB[32]. Private pharmacies are the preferable and first care unit which is approached by the patients in case of illness or disease. They also dispense the medications and prescriptions by private practitioners. Therefore, they are considered as crucial part of the health care provision chain. Pharmacists are aware of the diseases and symptoms and drug information for the treatment so they can play a vital role in referring the patients for tests[38].

## 5. RECOMMENDATIONS

It is recommended that the partnership model of public private sectors in TB case detection should be launched at larger scale in order to reduce the heavy TB burden in the Pakistan. This kind of public private collaborations needs strong support from the governmental institutions, in terms of supply of all the facilities and necessities for the better health of the people. Along with the facilities and financial support, government stewardship is required to make such projects successful on a larger scale. The Public private mix program in a country like Pakistan is necessary where most of the people prefer to visit a pharmacy instead of taking appointment from a doctor. This tendency towards increased case detection by involving private pharmacies indicates that the Public Private Mix program has a potential to improve case detection.On the basis of the findings of this study, it is recommended that this sort of programs in case of TB should be adopted on a larger scale involving private pharmacies as well as private hospitals and private physicians as they cover a larger population of people around the country.

## 6. LIMITATIONS OF THE STUDY

The study had a limitation. A few pharmacies did not participate in the study as they were not willing to participate without any incentives.

## 7. CONCLUSIONS

The study concludes that private pharmacies can be used as an effective first point contact to diagnose suspected patient. The role of pharmacies can be utilized to their full potential in early referral, diagnosis and treatment of TB cases. ThePPM model can be used effectively to detect TB cases timely. It can be extended and implemented in all provinces of Pakistanwith more technical and financial inputs in the existing system in order to completely eradicate TB and to enhance the role of Pharmacies in health care.

## Data Availability

All data produced in the present study are available upon reasonable request to the authors.

## LIST OF ABBREVIATIONS

CDR: Case Detection Rate
MOU: Memorandum of Understanding
NTP: National TB Control Program
PPM: Public Private Mix
TB: TB
WHO: World Health Organization

## DECLARATIONS

### Ethics approval and consent to participate

The study design was non-experimental and involved neither patient examination nor any intervention advised or made. All data collectors were required to sign a declaration assuring confidentiality and anonymity of data. A Memorandum of understanding (MOU) was signed with the universities and pharmacies which had relevant information regarding the project. Informed consents were obtained from all the participants of the study (who were randomly enrolled from the pharmacies). The study was regularly monitored and assessed throughout the program by project supervisors. The ethics approval was taken from the Ethics Committee on Human Research of University College of Pharmacy, University of Punjab with reference number HEC/PUCP/1953.

### Consent for publication

Not applicable.

### Availability of data and materials

The datasets used and/or analysed during the current study are available from the corresponding author on reasonable request.

### Competeing Interest

The authors declare that they have no competing interests.

## Acknowledgement

Authors acknowledge the contribution students of all pharmacy schools and staff members of pharmacies and medical stores.

